# Differential effects of illness and antipsychotics on cortical thinning in first episode psychosis: A randomised placebo-controlled MRI study

**DOI:** 10.1101/2025.05.08.25327204

**Authors:** Sidhant Chopra, Alexander Holmes, Ashlea Segal, Xi-Han Zhang, Shona M. Francey, Brian O’Donoghue, Vanessa Cropley, Barnaby Nelson, Jessica Graham, Lara Baldwin, Hok Pan Yuen, Kelly Allott, Mario Alvarez-Jimenez, Susy Harrigan, Avram J Holmes, Christos Pantelis, Stephen J Wood, Patrick McGorry, Alex Fornito

## Abstract

**Introduction:** Cortical grey matter loss is a common finding in Magnetic Resonance Imaging (MRI) studies of people with psychosis and has been shown to progress with ongoing illness. A major unresolved question concerns whether these changes are driven by the illness itself or represent iatrogenic effects of antipsychotic medication.

**Methods:** We report findings from a triple-blind randomised placebo-controlled MRI study where 62 antipsychotic-naïve people with first episode psychosis (FEP) received either an atypical antipsychotic or a placebo pill over a treatment period of 6 months. A healthy control group (n=27) was also recruited. Anatomical T1-weighed scans were collected at baseline, 3-months, and 12-months. Linear mixed effects models were fitted to >160,000 cortical surface loci to examine illness- and antipsychotic-related changes in cortical thickness. We also examined whether cortical changes were enriched within canonical functional networks or cytoarchitectonic types of laminar differentiation, and whether these changes spatially correlated with normative *in vivo* receptor/transporter densities via PET imaging and *ex vivo* transcriptomically-imputed cell densities.

**Results:** We found widespread cortical thinning over 12 months (p<.05_FDR_) in people with FEP who received placebo compared to healthy controls, with the largest effects occurring in frontal, cingulate, and occipital areas. No significant difference was detected in antipsychotic-treated people with FEP compared to healthy controls. Thickness differences in placebo group were not preferentially concentrated within any specific functional networks, although highly differentiated koniocortical areas were relatively protected from cortical thinning (p_spin_<.05_FDR_). Thinning in the placebo group spatially aligned with normative distributions of GABA_A/BZ_, 5HT_1B_, 5HT_2A_, and H_3_ receptors (.17 < r < .28; p_spin_<.05_FDR_). No significant associations between cortical thinning and symptom changes were identified.

**Conclusions:** Widespread thinning of cortical grey matter occurs over the first year of FEP in people who do not receive antipsychotic treatment. No such thinning is evident in patients receiving antipsychotics during the study intervention phase. These findings suggest that cortical thinning in the early stages of psychosis is an illness-related phenomenon that may be mitigated by early antipsychotic exposure.

---

Psychotic disorders represent a significant global health challenge, affecting approximately 24 million people^1^ worldwide and ranking in the top 20 causes of disability globally^2^. Decades of research and thousands of brain imaging studies have documented numerous quantitative brain differences in people with psychosis^3^, but we have thus far failed to uncover the specific causes of these brain changes.

A major impediment to uncovering such causes is the ubiquitous and near-immediate treatment of patients with antipsychotics, which makes it difficult to differentiate the effects of illness from medication on the brain. While longitudinal Magnetic Resonance Imaging (MRI) studies in patients who have received antipsychotics have documented progressive grey matter reductions^4–6^, the interpretation of such designs is limited by the confounding effect posed by covariations between medication exposure and illness severity.

Several lines of evidence suggest that both first-generation and second-generation antipsychotic medications may have neurotoxic properties. Primate and rodent studies have both demonstrated loss of brain volume, neurite, and glial density with prolonged exposure to antipsychotics^7–10^. Longitudinal MRI studies in patients have shown that volume reductions are often associated with cumulative antipsychotic exposure^11,12^ across both first and second generation medications^12,13^, with meta-analyses finding a similar association between exposure to first and second generation medications and brain structure at the study level^14–16^.

Other evidence supports a potentially neuroprotective effect of antipsychotic medications, particularly second-generation antipsychotics. Rodent studies have shown that some antipsychotic medications may interrupt pathological processes that drive illness-related structural brain changes^17^. Two large-scale naturalistic^4^ and controlled^18^ MRI studies have found that grey matter volume^18^ and cortical thickness^4^ reductions are attenuated in patients receiving second-generation compared to first-generation antipsychotics. These results are also consistent with a large body of clinical evidence showing that antipsychotics are effective in treating the acute symptoms of psychosis, improving quality of life, and reducing relapse and mortality^19–21^. However, some experimental studies have found that antipsychotic dose reduction during the early stages of psychosis may result in superior long-term recovery^22^ and that immediate introduction of antipsychotic medication may not be required for all cases to see functional improvement^23^.

Blinded, randomized, placebo-controlled MRI studies in first-episode psychosis (FEP) patients offer a gold standard design for disentangling whether structural brain changes in psychosis are caused by illness-related processes or antipsychotic exposure. Only two such studies have been conducted^13,24^. One demonstrated that maintaining olanzapine as an adjunctive therapy to sertraline was associated with significant cortical thinning compared to switching to placebo, in patients with psychotic major depressive disorder who had prior antipsychotic exposure^13^. In contrast, we recently found that volume loss in the basal ganglia during the first few months of illness observed in antipsychotic-naïve FEP patients is ameliorated patients administered second-generation antipsychotic medications^24^. We also found that changes in cortical grey matter volume were relatively minor in both medicated and unmedicated patients.

Grey matter volume may offer an insufficiently sensitive probe of cortical morphometry compared to cortical thickness. The latter can be measured with submillimeter precision^25^ and provides more precise information about underlying disease mechanisms than volumetric measures^26^, given that cortical surface area and thickness have distinct genetic origins^27–29^, developmental trajectories^30,31^, and biological substrates^32^.

Here, we combine the superior sensitivity of cortical thickness measures with our blinded, randomized, placebo-controlled design to disentangle the effects of illness and antipsychotic exposure on cortical anatomy within the first year of psychotic illness. We find widespread thinning in patients receiving placebo in the first 6 months of illness with no evidence for comparable changes in patients receiving second generation antipsychotics, suggesting that such medication may prevent or ameliorate illness-related cortical thinning within the first year of psychosis onset. We further show that the spatial pattern of illness-related cortical thinning is associated with specific molecular and cell-type distributions derived from independent PET imaging and independent post-mortem/ex vivo donor tissue, respectively, suggesting potential biological mechanisms of regional vulnerability to the neural effects of disease.

## Methods

### Participants

The study took place at the Early Psychosis Prevention and Intervention Centre, which is part of Orygen, Melbourne, Australia. Patients were aged 15–25 years and were experiencing a first episode of psychosis, defined as fulfilling Structured Clinical Interview for DSM-5 (SCID) criteria for a psychotic disorder, including schizophrenia, schizophreniform disorder, delusional disorder, brief psychotic disorder, major depressive disorder with psychotic symptoms, substance-induced psychotic disorder or psychosis not otherwise specified. Additional inclusion criteria designed to reduce clinical risk were: capacity to provide informed consent; English language comprehension; no MRI contraindications; duration of untreated psychosis (DUP) < 6 months; stable living arrangements; low self and interpersonal risk; and no or minimal prior antipsychotic medication exposure (<7 days of use or lifetime exposure equivalent to ≤1750 mg chlorpromazine; detailed specifications available in Supplementary Table 1). Our focus on people with FEP meant that, by definition, patients met criteria for multiple DSM-5 diagnostic categories. This transdiagnostic approach was adopted given that antipsychotic medication is a first-line treatment option across first-episode psychotic presentations.

Control participants were aged between 18-25 years, capable of providing written informed consent, and verified as psychiatrically healthy though a SCID and not at clinical high-risk through The Comprehensive Assessment of At-Risk Mental States (first four sub-scales). Control individuals currently using any psychotropic medications were excluded.

### Study Design

We conducted a triple-blind placebo-controlled longitudinal MRI study of antipsychotic medication. The current MRI study took place in the context of a larger clinical trial examining functional and clinical outcome^33,34^. Patients were randomized to one of two groups: one given second generation antipsychotic medication and the other given a placebo, with both patient groups receiving intensive psychosocial intervention. The third healthy control group received no intervention. Patients were assigned to treatment groups according to a stratified randomisation allocation, with Sex and DUP as factors. DUP was included as a three-level factor (0-30 days, 31-90 days, and >90 days; measured using clinical interview). Clinicians, patients, study assessors and researchers conducting MRI processing remained blinded to treatment allocation throughout the trial. For both patient groups, the treatment period spanned six months. MRI assessments were conducted at baseline, three months, and a final follow-up at 12 months. Supplement Figure 1 (SFig. 1) shows the recruitment flow diagram. Further research and safety protocols for the MRI study have been previously published^24^. The present analysis constitutes an ancillary analysis of the MRI data collected as part of a pre-registered clinical trial (ACTRN12607000608460; www.anzctr.org.au). Ethical approval for the study was granted by the Melbourne Health Human Research Ethics Committee (MHREC:2007.616).

### Antipsychotic medication

Patients randomised to the antipsychotic medication group received either 1mg risperidone (n=25) or 3mg paliperidone (n=5). To reflect real-world clinical treatment, this starting dose was then increased according to clinical response by the blinded treating clinician. The same procedure was followed for participants in the placebo group, who received a placebo pill that was identical in taste, appearance, and packaging to the active medication. Two individuals were exposed to antipsychotics at doses lower than the minimal exposure limit for inclusion in our study (<7 days of use or lifetime 1750 mg chlorpromazine equivalent exposure), and the remaining patients had no prior exposure.

Four patients within the placebo group were switched to open-label antipsychotic medication before the 3-month MRI scan and were excluded from the primary analysis. The randomization phase of the study terminated at 6 months, so patients in either treatment group could have received antipsychotic medication and ongoing psychosocial interventions in between the 6 and 12 months into the study. In practise, five patients in the placebo group were exposed to antipsychotic medication between the 6 and 12 month follow ups. To clearly disentangle the effects of illness from antipsychotics in our analyses, we only included patients in the placebo group who had no exposure to any antipsychotic medication throughout the 12 months of the study.

### Other medications and substance use

Substance use was measured using the World Health Organisation Alcohol, Smoking and Substance Involvement Screening Test (WHO-ASSIST). Concomitant medications were permitted during the trial, except for additional antipsychotics or mood stabilisers (rates of concomitant medication use are provided in Table 2).

**Table 1.**
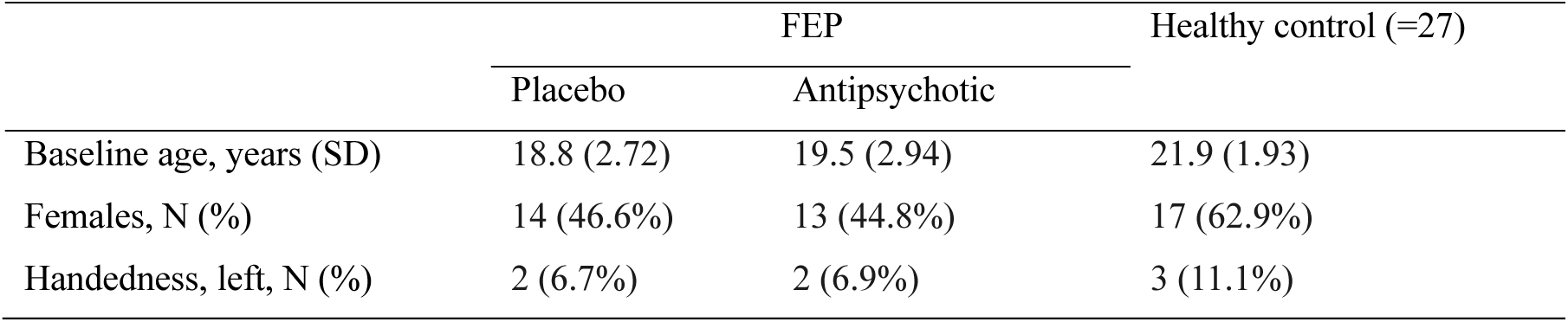
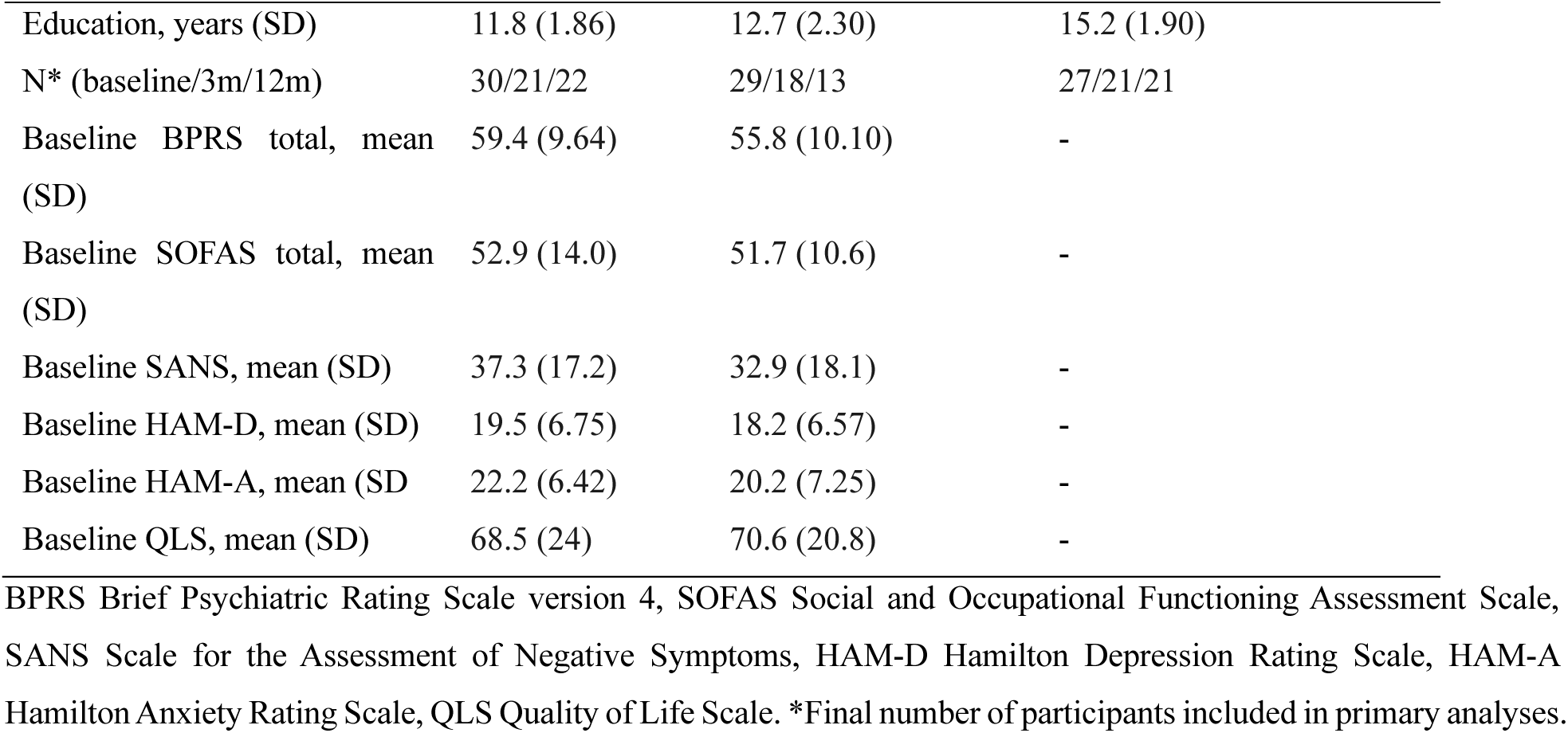
Sample characteristics.

**Table 2.**
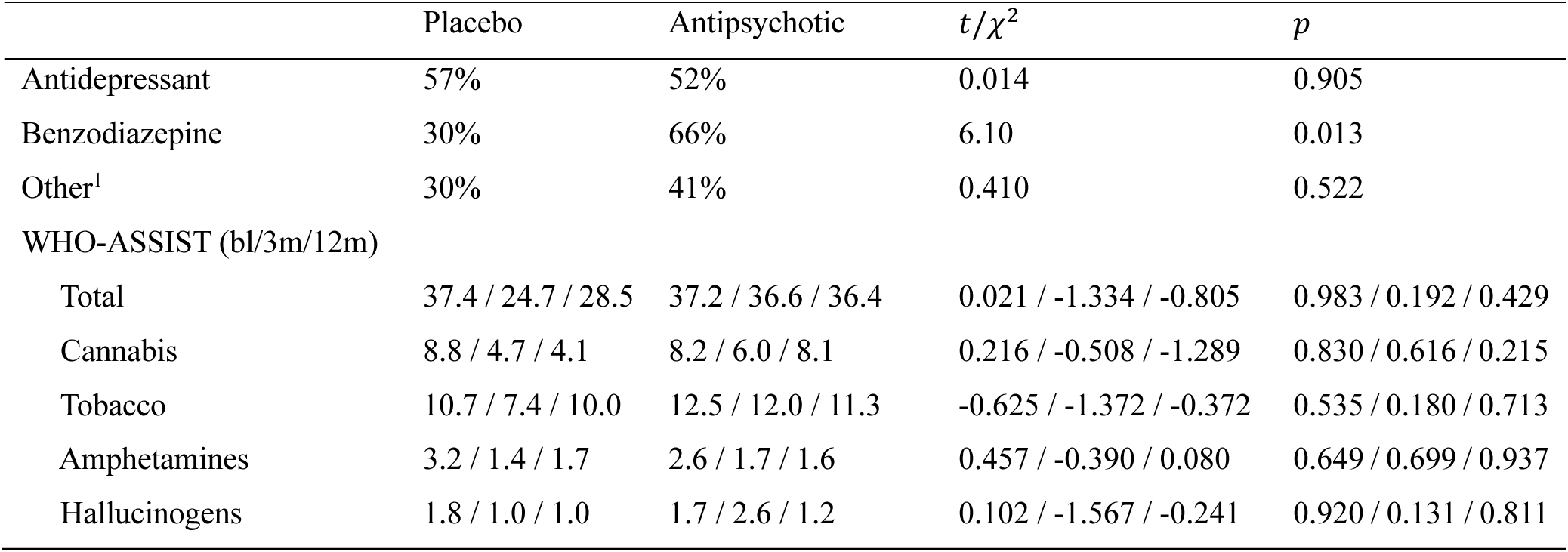
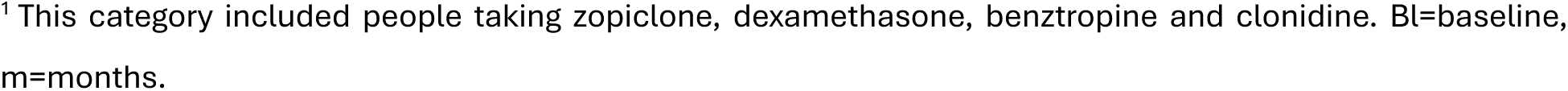
Concomitant medication and substance exposure.

| | Placebo | Antipsychotic | $t/\chi^2$ | $p$ |
| --- | --- | --- | --- | --- |
| Antidepressant | 57% | 52% | 0.014 | 0.905 |
| Benzodiazepine | 30% | 66% | 6.10 | 0.013 |
| Other<sup>1</sup> | 30% | 41% | 0.410 | 0.522 |
| WHO-ASSIST (bl/3m/12m) |  |  |  |  |
| Total | 37.4 / 24.7 / 28.5 | 37.2 / 36.6 / 36.4 | 0.021 / -1.334 / -0.805 | 0.983 / 0.192 / 0.429 |
| Cannabis | 8.8 / 4.7 / 4.1 | 8.2 / 6.0 / 8.1 | 0.216 / -0.508 / -1.289 | 0.830 / 0.616 / 0.215 |
| Tobacco | 10.7 / 7.4 / 10.0 | 12.5 / 12.0 / 11.3 | -0.625 / -1.372 / -0.372 | 0.535 / 0.180 / 0.713 |
| Amphetamines | 3.2 / 1.4 / 1.7 | 2.6 / 1.7 / 1.6 | 0.457 / -0.390 / 0.080 | 0.649 / 0.699 / 0.937 |
| Hallucinogens | 1.8 / 1.0 / 1.0 | 1.7 / 2.6 / 1.2 | 0.102 / -1.567 / -0.241 | 0.920 / 0.131 / 0.811 |

<sup>1</sup> This category included people taking zopiclone, dexamethasone, benztropine and clonidine. Bl=baseline, m=months.

### MRI acquisition and processing

A 3-Tesla Siemens Trio Tim scanner located at the Royal Children’s Hospital in Melbourne, Australia, was used to acquire a high resolution structural T1-weighted scan for each participant at each timepoint. For each scan, a total of 176 slices were acquired with an interleaved acquisition using the following parameters: TR = 2300ms; TE = 2.98ms; flip angle of 9°; FOV of 256mm; voxel size of 1.1 x 1.1. x 1.2 mm.

Following visual quality control of raw T1-weighted data, we implemented the FreeSurfer version 6.0 longitudinal (Martinos Center for Biomedical Imaging) processing pipeline^35^, using a within-participant template estimation for unbiased longitudinal analysis^36^. The software reconstructs the inner and outer cortical surfaces using a triangular mesh. To minimise errors in the delineation of the pial and white matter surfaces, boundary errors were manually corrected for all scans as per previous guidelines (https://surfer.nmr.mgh.harvard.edu/fswiki/LongitudinalEdits). Individual-level thickness maps were resampled from native subject space to a common space comprising 163,842 vertices per hemisphere (*fsaverage*) for group-level analyses. Consistent with prior recommendations^37^, a 15-mm full-width half maximum Gaussian smoothing kernel was applied to increase the signal-to-noise ratio and improve spatial registration while preserving true regional effects. To evaluate the effect of spatial smoothing kernel size on our primary results, we also report findings using 10 mm and 20 mm kernels (see SFig2).

### Cortical thickness analysis

Mass-univariate linear mixed effects models were implemented to examine cortex-wide cortical thickness changes. The FreeSurfer MATLAB toolbox^37,38^ was used to build spatiotemporal linear mixed effects models fitted to each of 327,684 vertices comprising the cortical surface meshes of the left and right hemispheres. Each model included time from first scan (days), group, group by time interaction, age at baseline and sex as fixed effects and subject as a random subject-specific intercept and slope. The statistical significance of the group by time interaction (F-statistic) was assessed at p < .05, using a two-stage False Discovery Rate (FDR) procedure^39^ for multiple comparison correction. To examine cortical thickness changes, we used two key contrasts, consistent with our previous work in this cohort^40^. First, we isolated illness-related effects by comparing changes over time between the placebo and healthy control groups. Second, we mapped antipsychotic-related effects by comparing changes in the antipsychotic medication group against the combined changes in the placebo and healthy control groups. Given that inclusion of the placebo group in the contrast examining antipsychotic-related effects may obscure changes in the medication group, we also conducted a secondary contrast comparing the antipsychotic medication group to just the healthy control group (SFig. 4). These contrasts were conducted at two intervals: baseline to 3 months and baseline to 12 months, with the latter analysis examining linear changes across all three timepoints.

### Symptom and functional associations

To assess whether changes in cortical thickness are associated with changes in functioning and symptoms, we adapted a cross-validated predictive modelling framework^41,42^. First, we computed proportional change scores at each vertex for patients who had complete clinical and imaging data from baseline to 3-months (Δ3-months) and baseline to 12-months (Δ12-months). We then computed proportional change scores for the two preregistered primary trial outcome measures^43^––total scores for the Social and Occupational Functioning Assessment Scale and the Brief Psychiatric Assessment Scale––at each of two longitudinal timepoints (Δ3-months, Δ12-months). For Δ3-months and Δ12-months analysis, 38 and 34 patients with complete data at both timepoints, respectively, were included in the analysis. We implemented predictive modelling using a nested *k*-fold cross-validation procedure. This involved repeatedly dividing participants into four folds, with three folds used for training and one held-out fold for testing. The training process involved selecting features (vertices) based on partial correlations between volume change and behavioural change scores (p < .01, uncorrected) white adjusting for covariates (age, sex, cumulative antipsychotic exposure) and then summing the weights of positively and negatively correlated edges separately, resulting in two condensed summary features per participant. These summary features were used to train general linear models that predicted behavioural changes in the held-out test set. The product-moment correlation between observed and predicted change score within the held-out test set was used to assess prediction performance.

The entire *k*-fold modelling process was repeated 100 times to estimate the sampling error of the model’s prediction performance. To assess model significance, we permuted the behaviour scores 1000 times and repeated the *k*-fold modelling process, resulting in a null distribution of product-moment correlations. We then estimated *p* values as the proportion of null correlations greater than the mean observed model performance across the 100 repeated runs of the *k*-fold modelling process *r*_mean_. Further details on modelling and exploratory secondary outcome measures can be found in the Supplement.

### Enrichment analyses

To understand the potential neural substrate of any detected cortical changes, we examined whether thickness differences were preferentially enriched within certain functional networks, cytoarchitectonic types, or correlated with normative distributions of key neurotransmitters or cell-type distributions inferred using transcriptomic data. To this end, vertex-level t-statistic maps were first parcellated into 300 discrete cortical areas^44,45^ using a previously validated atlas. Enrichment relative to canonical functional networks was evaluated with respect to the 7 intrinsic functional networks defined by Yeo and Krienen^44,45^. Enrichment relative to cytoarchitectonic types was assessed relative to the von Economo and Koskinas atlas^46–48^ using a winner-take-all classification^47,48^, to rank different cortical areas according to their level of laminar differentiation. Under this scheme, primary unimodal areas tend to show higher laminar differentiation than transmodal and limbic regions. Enrichment in both cases was quantified by averaging the *t*-statistic for cortical thickness differences in each area of each network/cytoarchitectonic type (Fig2A-B) and comparing the magnitude of these regional averages to those obtained under a spatial spin test^48,49^ that involves rotating the underlying map of cortical thickness differences in random directions. The resulting null map thus preserves lower-order spatial effects of no interest, such as spatial autocorrelation, while randomizing the spatial association of interest. We used 10,000 rotations to generate the null distributions. Statistical significance was assessed at p<.05 (two-tailed) with FDR correction applied to account for the number of different networks and subdivision.

**Fig1.**
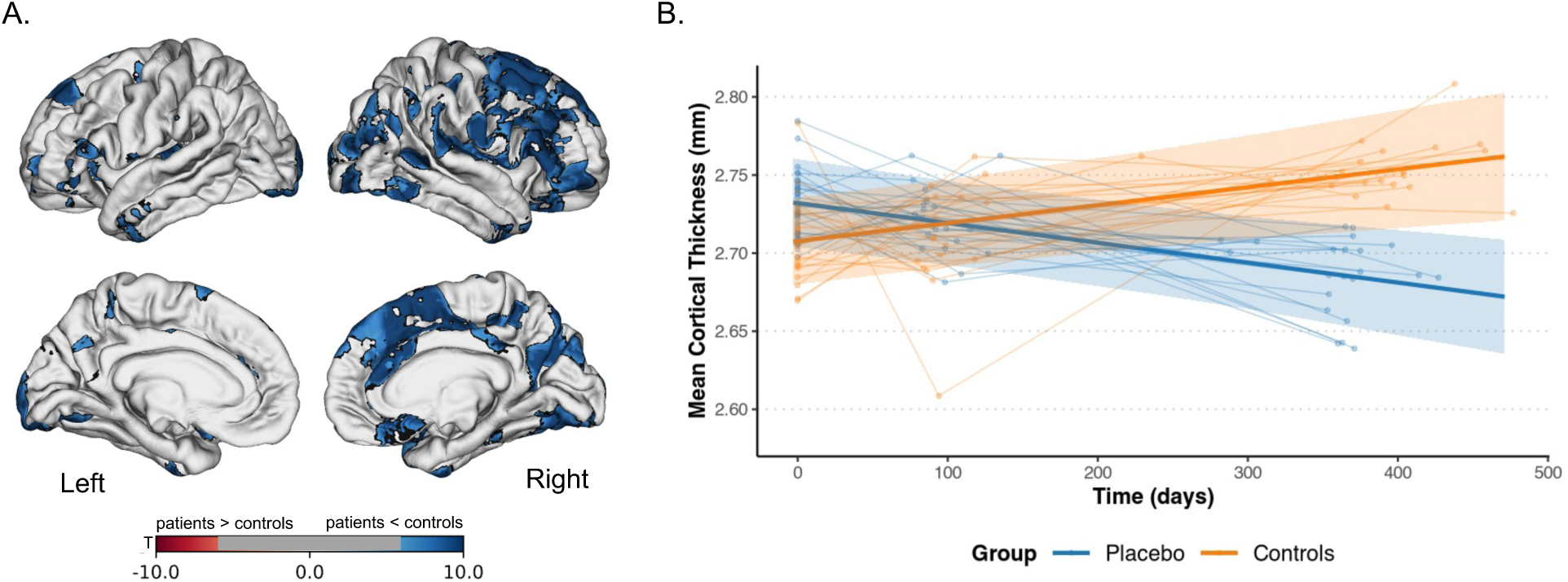
Illness-related cortical thinning. A) Cortex-wide effect-size map of False-Discovery Rate (FDR) corrected (p_FDR_<.05) cortical thickness changes in the placebo patient group comparted to healthy control group from baseline to 12months. Blue indicated thinning in the patient group compared to controls, whereas red indicates increases. B) Group-level marginal effects from a linear mixed-effects model fit to the mean cortical thickness of significant vertices, overlayed over the covariate adjusted, subject-level mean cortical thickness.

**Fig 2.**
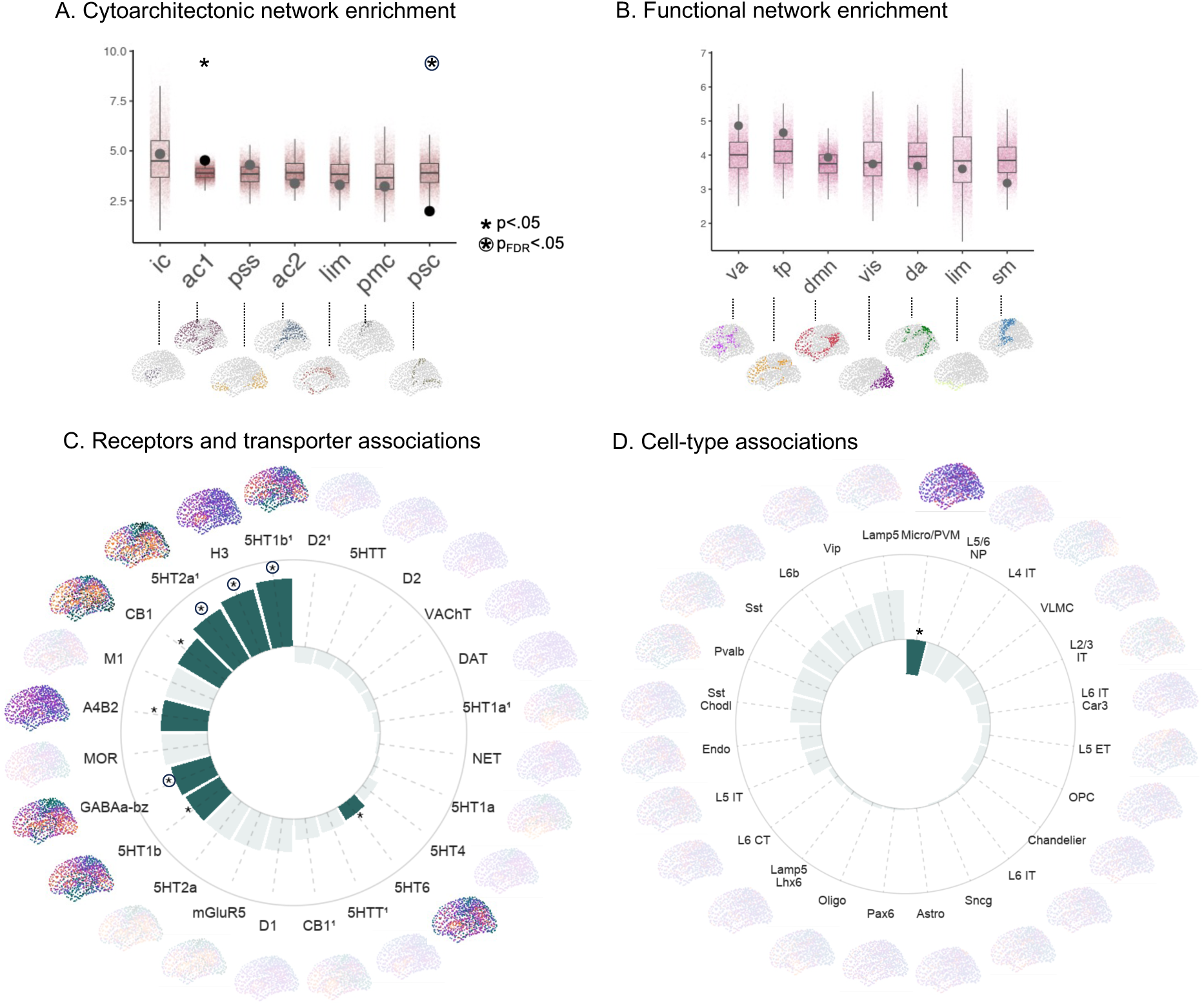
Network, neurochemical and cellular correlates of illness-related cortical thinning. **(A-B)** Black dots represent observed mean test-statistic of illness-related cortical thinning in patients, with coloured dots representing null mean test-statistic generated using spatial-autocorrelation preserving models, for each of 7 cytoarchitectonic types **(A)** defined according to the von Economo and Koskinas atlas^46–48^, and **(B)** 7 canonical functional networks defined by Yeo and Krienen^44,45^. **(C-D)** Univariate associations between illness-related cortical thinning and normative receptor/transporter distributions measured *in vivo* using PET imaging **(C)**, and 23 transcriptomically-imputed cell densities measured *ex vivo **(D)***. Asterisks (*) Indicates statistical significance (p<.05), with a circle indicating survival against False-discovery rate (FDR) correction. pss=primary/secondary sensory cortex, psc=primary sensory cortex, lim=limbic cortex, ac1=association cortex 1, ac2= association cortex 2, ic=insular cortex, pmc=primary motor cortex. dmn=Default-mode, vis=Visual, fp=Frontoparietal, va=Ventral attention, sm=somatomotor, lim=limbic, da=Dorsal attention.

The correspondence between cortical thickness changes and normative neurochemical receptor and cellular densities was assessed using two sources of data. Normative neurochemical densities were obtained from 24 different group-consensus PET binding maps of receptor and transporter densities, covering aspects of serotonin, glutamate, adrenergic, GABA, dopamine, opioid, cannabinoid and cholinergic systems from healthy populations, as collated in the *neuromaps* toolbox^50^. For PET tracers that had multiple datasets, a sample-size weighted average map was computed (Fig2). Each PET map was parcellated into the same 300 region atlas as previously described.

Regional transcriptomic cell-type concentrations were deconvolved from whole brain bulk tissue transcriptions provided by the Allens Human Brain Atlas^51^, with 24 different cell-types being imputed using recently published single-nucleus RNA-sequencing data^52^ (see Zhang et al^53^ for detailed overview). Briefly, the transcriptional signatures identifying each cell class in the Allen Human Brain atlas bulk samples were derived from cortical single-nucleus RNA sequencing (snRNA-seq) data of eight cortical areas. This included 24 cellular classes with distinct laminar specialization, developmental origin, morphology, spiking pattern, and broad projection targets^54^. These cells include 9 GABAergic inhibitory interneurons (PAX6, SNCG, VIP, LAMP5, LAMP5 LHX6, Chandelier, PVALB, SST CHODL, Sst), 9 glutamatergic excitatory neurons (L2/3 IT, L4 IT, L5 IT, L6 IT, L5 ET, L5/6 NP, L6 CT, L6b, L6 IT Car3), and 6 non-neuronal cells (Astro, Endo, VLMC, Oligo, OPC, Micro/PVM). This procedure resulted in region-level cell density estimates for each of the 24 cell-types. Associations between regional cortical thickness change maps and normative neurochemical or cell density maps were assessed using a product-moment correlation (*r*). Statistical significance was assessed at p<.05 (two-tailed) using the previously described spin-test with 10,000 permutations to generate null distributions of the spatial correlations. FDR correction was implemented to account for the number of receptors/transporters or cell-types.

## Results

### Demographics and clinical characteristics

We have previously reported the demographics and clinical characteristics of this cohort^23^. Briefly, there were no significant differences between the patient and control samples in sex or handedness, but the patients were, on average, 1.9 years younger (t=−4.49; p < .001) and had 2 years less education (t=−6.21; p < .001; see Table 1 for further details). Baseline diagnostic breakdown is provided in STable1.

### Cortical thinning in antipsychotic-naïve patients

We detected a significant group by time interaction (p<.05_FDR_) across 62,043 vertices, showing wide-spread and prominent cortical thinning in patients who received placebo compared to healthy control participants over 12-months (Fig1). Across significant vertices, patients receiving placebo showed a decrease of 2.21% (95% CI: 0.60% - 3.80%), whereas control participants showed an increase of 1.25% (95% CI: 0.91% - 3.00%). Cortical thinning over 12-months in the placebo group was especially apparent in bilateral pre-frontal, anterior temporal, and occipital areas, as well as in right orbitofrontal, medial frontal, cingulate and parietal areas (Fig1A). No significant effects were detected over 3-months (SFig3). No significant changes in cortical thickness were detected over 3-months or 12-months in the antipsychotic group comparted to both healthy control and placebo groups (SFig3), nor when compared to the healthy control group alone (SFig4). No changes in surface area were detected for either group. No significant associations between change in thickness and change in symptoms or functional outcomes were detected at 3- or 12-months (SFig5).

### Potential confounds

We found a significant difference between the placebo and antipsychotic medication groups on benzodiazepine exposure during the treatment period (*χ*^2^ = 6.10; p=0.013). We re-computed all longitudinal analyses examining cortical thickness after including benzodiazepine exposure as a covariate and similar results (SFig6). There were no significant differences between the treatment groups with respect to antidepressant or other medications exposure or substance use.

### Enrichment analyses

Cortical thinning over 12-months in the placebo group showed no preferential enrichment within any of the Yeo and Krienen^44^ canonical functional networks. We found some evidence for an enrichment within cytoarchitectonically defined association cortex coupled with a relative sparing of early maturing Koniocortical/primary sensory areas, but only the latter survived multiple comparison correction (p_FDR_<.05). Cortical thinning over 12 months in the placebo group was also spatially correlated with normative distributions of GABA_A/BZ_, 5HT_1B_, 5HT_2A_, and H_3_ receptors (.17 < r < .28; p_FDR_ <.05; Fig3A). A suggestive association with microglia/perivascular macrophage density was identified, but did not pass multiple comparison correction (r=-.103; p=.004; Fig3B). No other cell types were implicated.

## Discussion

The primary aim of this study was to understand whether cortical thinning in antipsychotic-naïve people experiencing a first episode of psychosis (FEP) is driven by the illness itself or represents iatrogenic effects of antipsychotic medication. Using a randomized placebo-controlled longitudinal design, we found that patients receiving placebo demonstrated widespread cortical thinning compared to healthy controls. No significant cortical thickness differences were detected in patients who received antipsychotics, nor over the first three months in either patient group. These findings are consistent with the view that cortical thinning in early illness stages cannot be attributed to antipsychotic treatment and that is not prevented by psychosocial treatment alone. Indeed, our results suggest that antipsychotic treatment may prevent or ameliorate illness-related cortical thinning. Notably, areas high in GABA_A/BZ_, 5HT_1B_, 5HT_2A_, and H_3_ receptors appear particularly vulnerable whereas early maturing and highly differentiated primary unimodal cortices are relatively spared from these illness-related effects.

### Cortical thinning in the absence of antipsychotic medication

The widespread use of antipsychotic medications during the early stages of psychosis has made it difficult to disentangle whether these medications drive or prevent the loss of brain tissue. Our analysis indicates that antipsychotic-naïve patients who do not receive antipsychotics during the first 6 months of treatment engagement show widespread cortical thinning that is particularly prominent in bilateral prefrontal, anterior temporal, and occipital areas, as well as in right orbitofrontal, medial frontal, cingulate and parietal areas. These results are consistent with a large body of cross-sectional MRI studies showing a thinner cortex across different stages of psychosis^55^, post-mortem studies showing reductions in brain weight^56^, and longitudinal MRI studies in patients showing accelerated thinning during the first few years of illness^57–60^. Studies in high-risk patients who convert to psychotic illness have now consistently demonstrated accelerated thinning of the prefrontal cortex^58,61,62^. Our findings extend this work to show that accelerated thinning may continue during the first year and encompasses widespread cortical areas in patients who are not exposed to antipsychotic medications.

While our findings are consistent with an illness-related neuroprogressive hypothesis of schizophrenia, the term ‘illness-related’ should not simply ascribed to a direct pathophysiological effect of psychosis. Rather, the cortical thinning observed may reflect a confluence of factors, including secondary effects related to systemic inflammation, sedentary lifestyle, dietary influences, and chronic stress^63,64^, or reflect compensatory mechanisms, such as adaptive synaptic refinement^65^. In fact, several studies have reported that cortical thinning in psychotic illness may, in certain contexts, be associated with a more favourable pattern of functional activation^66–68^. Thus, while tissue loss remains a hallmark of the illness and has been linked with greater symptom severity^24,69^, it is essential to recognize that illness-related brain changes measured using MRI likely represent a multifaceted process. Moreover, it is important to emphasize that the high rates of cortical thinning observed here likely do not persist linearly over time. A substantial body of evidence indicates that brain changes in psychosis follow a non-linear trajectory^55^, with some neuroanatomical deviations attenuating at later stages of illness^70^. Future longitudinal studies integrating multimodal imaging with clinical control groups and detailed assessments of lifestyle and biological stressors will be crucial for disentangling these complex mechanisms and refining our understanding of the prognostic implications of cortical thinning in psychosis.

### Does antipsychotic exposure prevent cortical thinning?

We did not find any statistically significant cortical thinning in the group of patients that received antipsychotic medications. This result is consistent with findings in other medication exposed early-stage psychosis cohorts showing a lack of structural loss^71–74^. Given the thinning seen in the placebo group, our findings suggest a protective effect of antipsychotics. While other longitudinal studies and meta-analyses have demonstrated that a higher cumulative dose of antipsychotic medication is associated with greater levels of tissue loss^11,12,14–16^, these studies have been confounded by the fact that patients with more severe symptoms are typically given higher antipsychotic doses.

Our findings in the medicated group contrast with other studies that have demonstrated statistically significant cortical thinning in medication-exposed first-episode cohorts^55,75^. This discrepancy may be due to power, sample size or distinct characteristics of our sample and study methodology, such as the intensive psychosocial treatment provided to all patients (also see *Limitations*). Alternatively, recent work has suggested that the putative antipsychotic-related tissue loss reported in extant studies may be related to lack of medication adherence^76,77^. Controlled studies such as ours, as well as studies using long-acting injectable antipsychotics—where medication adherence and cumulative exposure can be precisely measured—have demonstrated that the observed structural loss is not dose-dependent and is less likely to occur when patients are verifiably taking antipsychotics^77–80^.

Our findings are in line with large-scale longitudinal MRI studies that have demonstrated attenuated cortical tissue loss in patients assigned to second-generation (as opposed to first-generation) antipsychotics such as those used in the current study^18,81,82^ This finding is also consistent with *in vitro* and animal models demonstrating protective effects of second-generation antipsychotics, potentially related to several candidate mechanisms, including neurogenesis^17^ and protection against oxidative stress^83,84^. However, MRI is unable to identify a specific cellular mechanism that would verify a neuroprotection hypothesis, and the molecular mechanisms by which second-generation medications might protect brain tissue in humans are still poorly understood. It is possible any protective effects of antipsychotics on brain tissue occur as a secondary consequence of behavioural changes associated with improvement in symptoms, quality of life and functioning^85^. However, while patients in the medication group did show significantly lower negative symptoms at 12 months compared to the placebo group, no differences in any other clinical and functional outcome were found in the current trial^23^.

### Cellular and molecular correlates of cortical thinning

Our enrichment analyses indicated that highly differentiated koniocortical areas are relatively protected from thinning. These primary sensory (visual, auditory, and somatosensory) areas are defined by well-developed layers II and IV, containing dense populations of small granule neurons with short local axons^86^ and are also among the earliest cortical areas to mature during development^31^. This contrasts with later-maturing higher-order transmodal association areas, which a large body of work has demonstrated as major sites of pathology in psychosis^87^. Our findings suggest that the accelerated maturation of koniocortical areas and their distinctive cellular and molecular features may confer resilience to the effects of psychosis.

We also found that areas of progressive thinning are associated with the normative spatial distributions of several neurotransmitter receptor systems: 5HT_1B_, 5HT_2A_, H_3_ and GABA_A/BZ_. The spatial colocalization of these receptor gradients with regions of thinning suggests that receptor-mediated mechanisms may underlie regional vulnerability to cortical thinning. Both *in vivo*^88^ and *ex vivo*^89,90^ studies of people with psychosis have reported altered levels of monoaminergic receptors. Findings regarding GABAergic receptor densities are mixed^91^, but aberrant GABAergic cortical interneuron structure and function remains one of the most widely replicated postmortem and genetic observations^92,93^. Second-generation antipsychotics, which possess a high affinity for 5-HT₂_A_ receptors, have been shown to increase neurotrophic factors, thereby promoting neuroplasticity and facilitating the repair of dendritic spines^17^. This raises the possibility that the engagement of 5-5HT_2A_ receptors could counteract thinning by promoting synaptic repair and potentially stabilizing cortical volume. Moreover, although data on GABAergic modulation are less consistent^94,95^, it is plausible that antipsychotics may also influence GABA-mediated signalling pathways, potentially restoring excitatory/inhibitory balance and contributing to cortical preservation^96^. Further mechanistic studies, integrating molecular imaging with longitudinal anatomical assessments, will be crucial to understand the precise receptor-specific pathways that contribute to structural preservation.

### Limitations

The practical challenges of recruiting medication-naïve individuals limited the sample size of the current cohort. Our strict inclusion criteria (i.e., low risk of harm to self or others, lived in stable accommodation, and short DUP) raise concerns that our sample comprises patients with a mild form of illness. This would suggest that our findings in Fig 1 offer an optimistic indication of the extent of brain changes observed in more severe patients. This seems unlikely, however, as the mean baseline BPRS score of patients in our study would classify them as markedly ill^97^ and the mean baseline SOFAS score was consistent with serious functional impairment (see Chopra et al.^24^ for a detailed discussion on generalizability). We only examined risperidone and paliperidone and it is therefore unclear whether our results generalise to other antipsychotic medications.

## Conclusions

We observed widespread cortical thinning in people with FEP receiving placebo compared to healthy controls that was not apparent in patients exposed to antipsychotics from illness onset. Our findings indicate that longitudinal cortical thinning in early psychosis cannot be attributed to antipsychotic medications and that treatment with such agents may prevent illness-related cortical tissue loss during a first episode. Serotonergic, histaminergic, and GABAergic systems may influence regional vulnerabilities to the cortical effects of psychotic illness.

## Data Availability

Subject-level produced in the present study will available upon appropriate ethics approval and reasonable request to the authors. Group-level de-identified data will be made available upon publication.

## Funding and disclosures

Janssen-Cilag partially supported the early years of this study with an unrestricted investigator-initiated grant and provided risperidone, paliperidone, and matched placebo for the first 30 participants. The study was then funded by an Australian National Health and Medical Research Project (NHMRC) grant (Grant No. 1064704 [to SMF, BO, BN, HPY, KA, MA-J, SH, SJW, PM, AF]). The funders had no role in study design, data collection, data analysis, data interpretation, writing, approval or submission of this manuscript. C Pantelis was supported by a National Health and Medical Research Council L3 Investigator Grant (1196508). AF was supported by the NHMRC (ID: 1197431), Australian Research Council (ID: FL220100184), and Sylvia and Charles Viertel Charitable Foundation.

This study has been supported by a large number of clinical staff at Orygen Youth Health (Craig Macneil, Kingsley Crisp, Dylan Alexander, Tina Proffitt, Rachel Tindall, Jennifer Hall, Lisa Rumney, Franco Scalzo, Melissa Pane, Linda Kader, Frank Hughes, Clare Shelton, Ryan Kaplan, David Hallford, Bridget Moller, and Rick Fraser) and research assistants (Daniela Cagliarini, Suzanne Wiltink, Janine Ward, and Sumudu Mallawaarachichi). The trial took place at the Early Psychosis Prevention and Intervention Centre, which is part of Orygen Youth Health, Melbourne, Australia.

Data processing was conducted using Multimodal Australian ScienceS Imaging and Visualisation Environment (Goscinski, 2014).

## Supplement

**SFig. 1.**
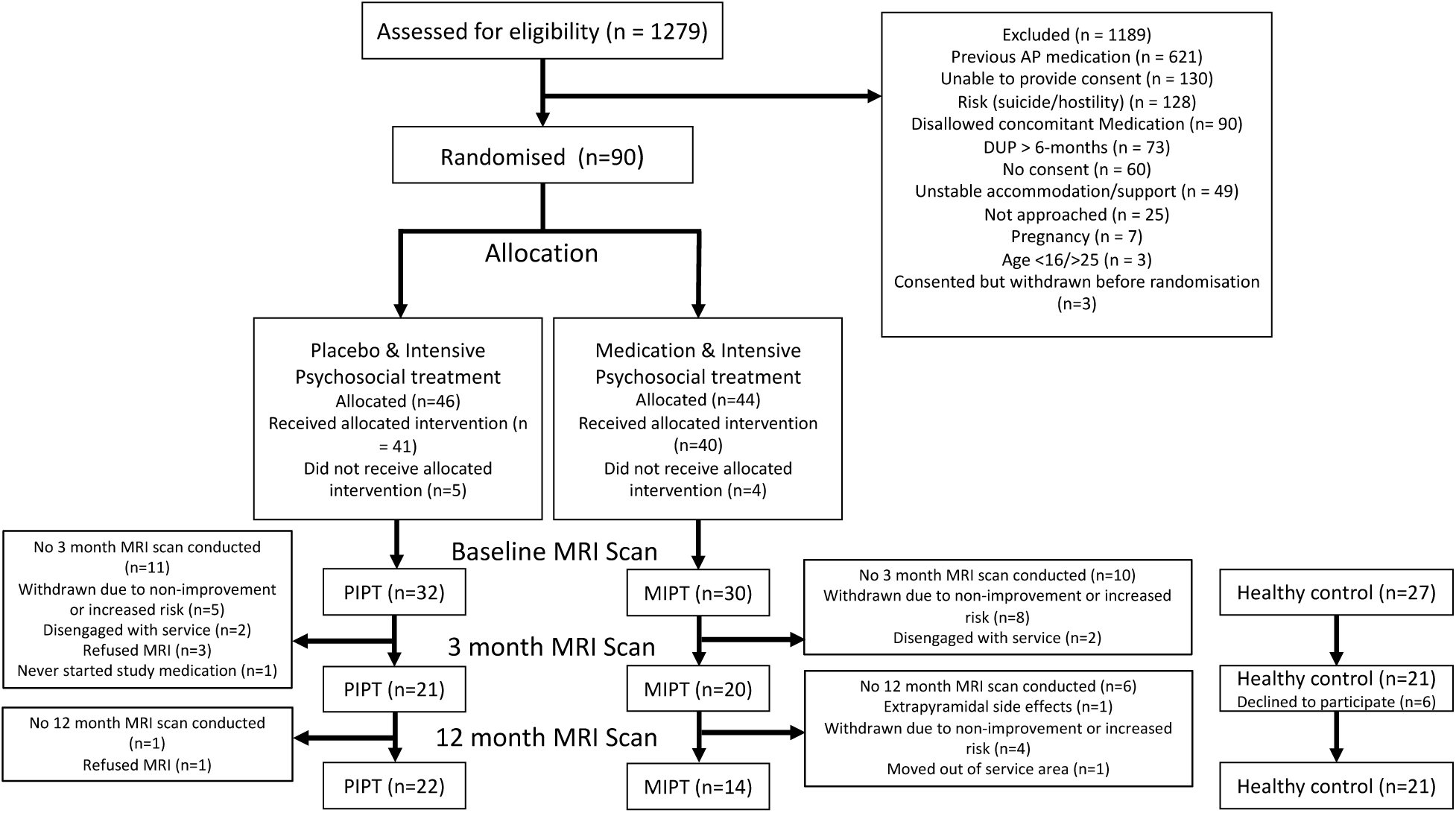
The flow of patients and healthy control participants though the study. Previously published in Chopra et al., (2019)^24^.

**SFig2.**
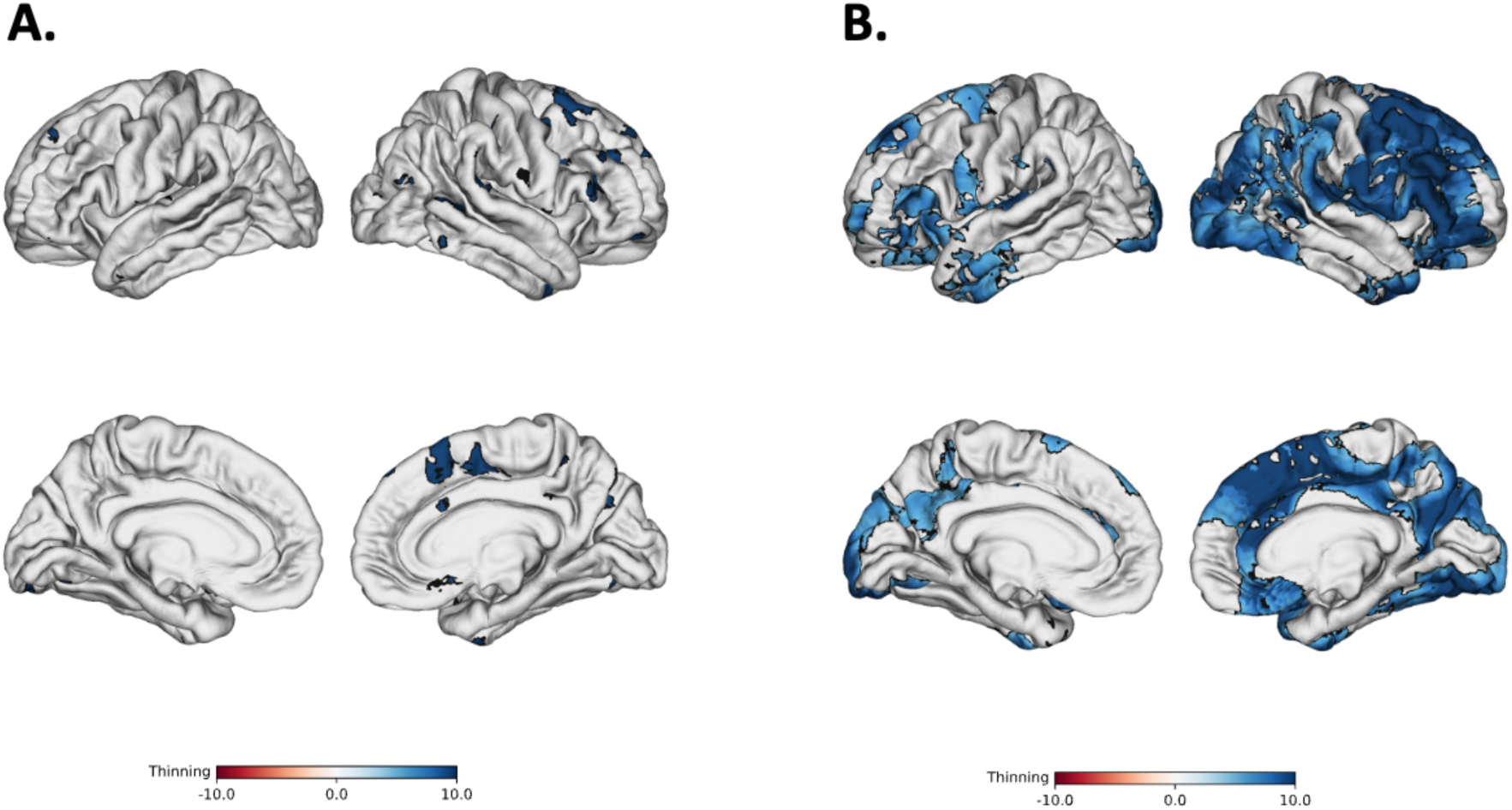
Illness-related cortical thinning using alternate smoothing kernels. Cortex-wide effect-size map of False-Discovery Rate (FDR) corrected (p_FDR_<.05) cortical thickness changes in the placebo patient group comparted to healthy control group from baseline to 12months using 10mm **(A)** and 20mm **(B)** alternate smoothing kernels.

**SFig3.**
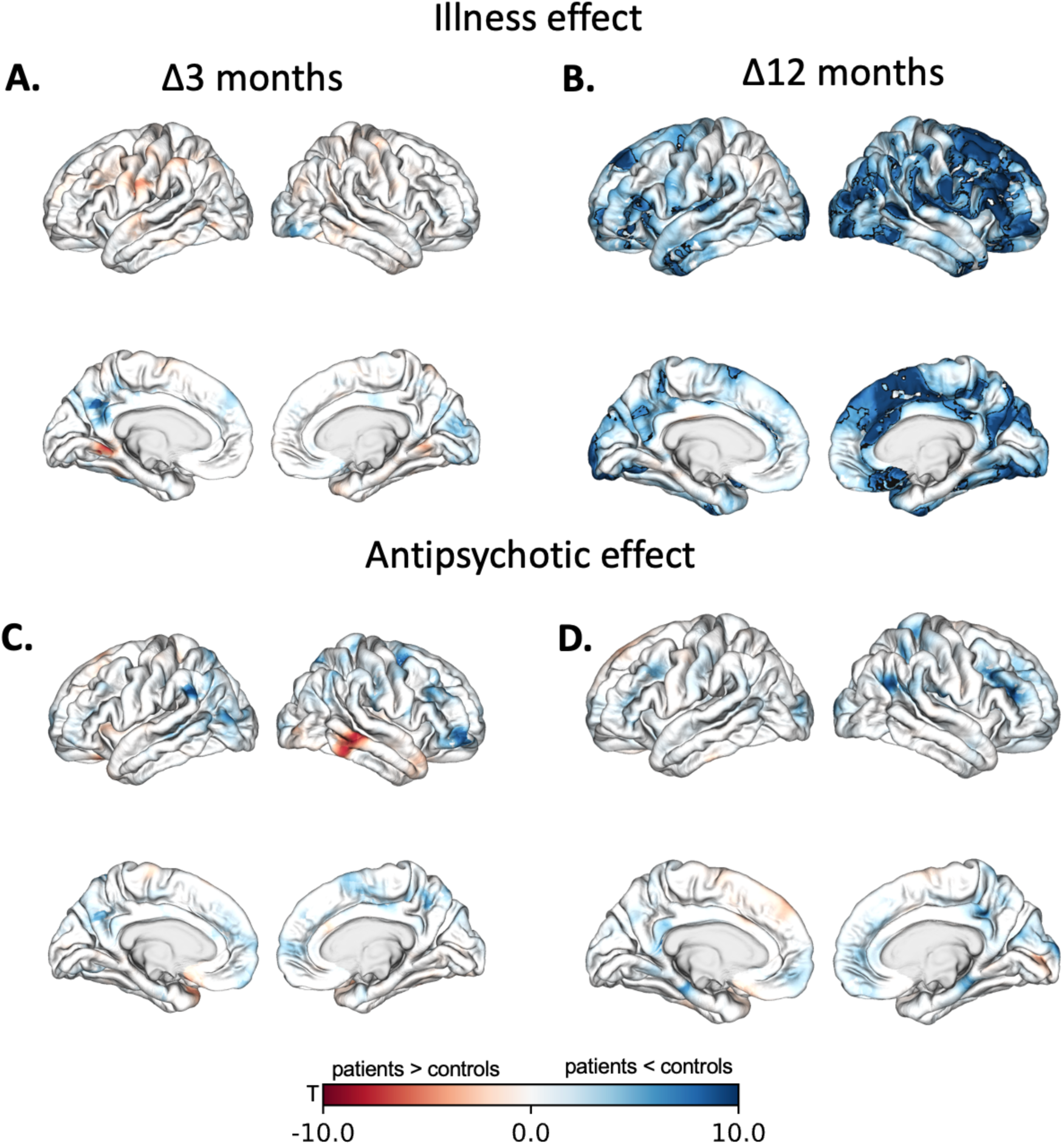
Unthrehsolded cortical thickness results. **A-B)** Show cortex-wide test statistic maps for illness-related cortical thickness changes (Placebo group vs. Healthy Controls) from baseline to 3-months **(A)** and baseline to 12-months **(B)**. Statistically significant effects (p<.05_FDR_) are outlines in black were only found in at baseline to 12-months (also displayed in Fig1). **C-D)** Show cortex-wide test statistic maps for antipsychotic-related cortical thickness changes (Antipsychotic group vs. Healthy Controls & Placebo groups) from baseline to 3-months **(C)** and baseline to 12-months **(D)**. No Statistically significant effects (p<.05_FDR_) were detected.

**SFig4.**
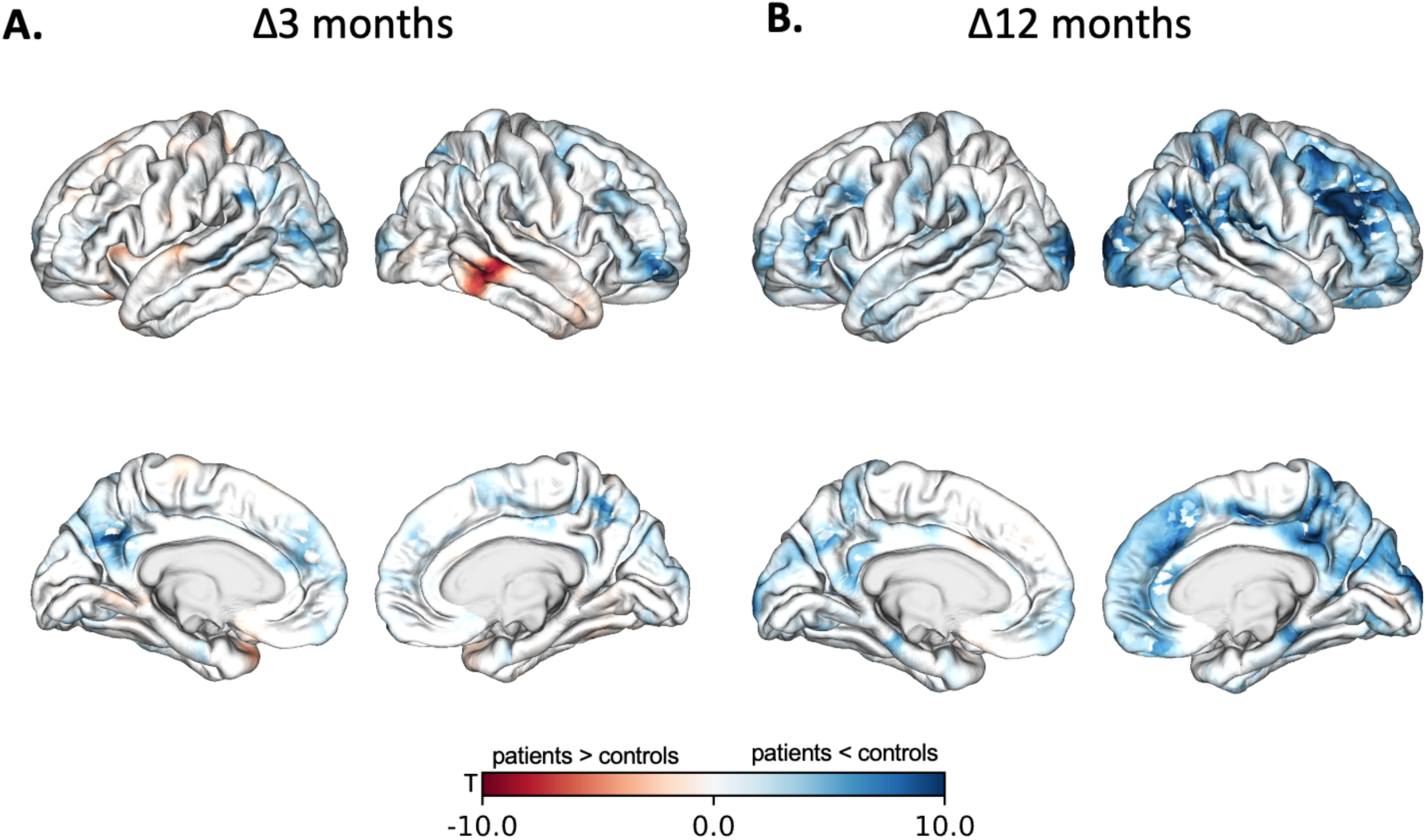
Unthrehsolded cortical thickness results for a contrast comparing change in cortical thickness in the antipsychotic group to only the healthy controls at baseline to 3-months **(A)** and baseline to 12-months **(B)**. As with the antipsychotic-related effects shown in SFig2, no significant effects (p < .05_FDR_) were detected.

**SFig5.**
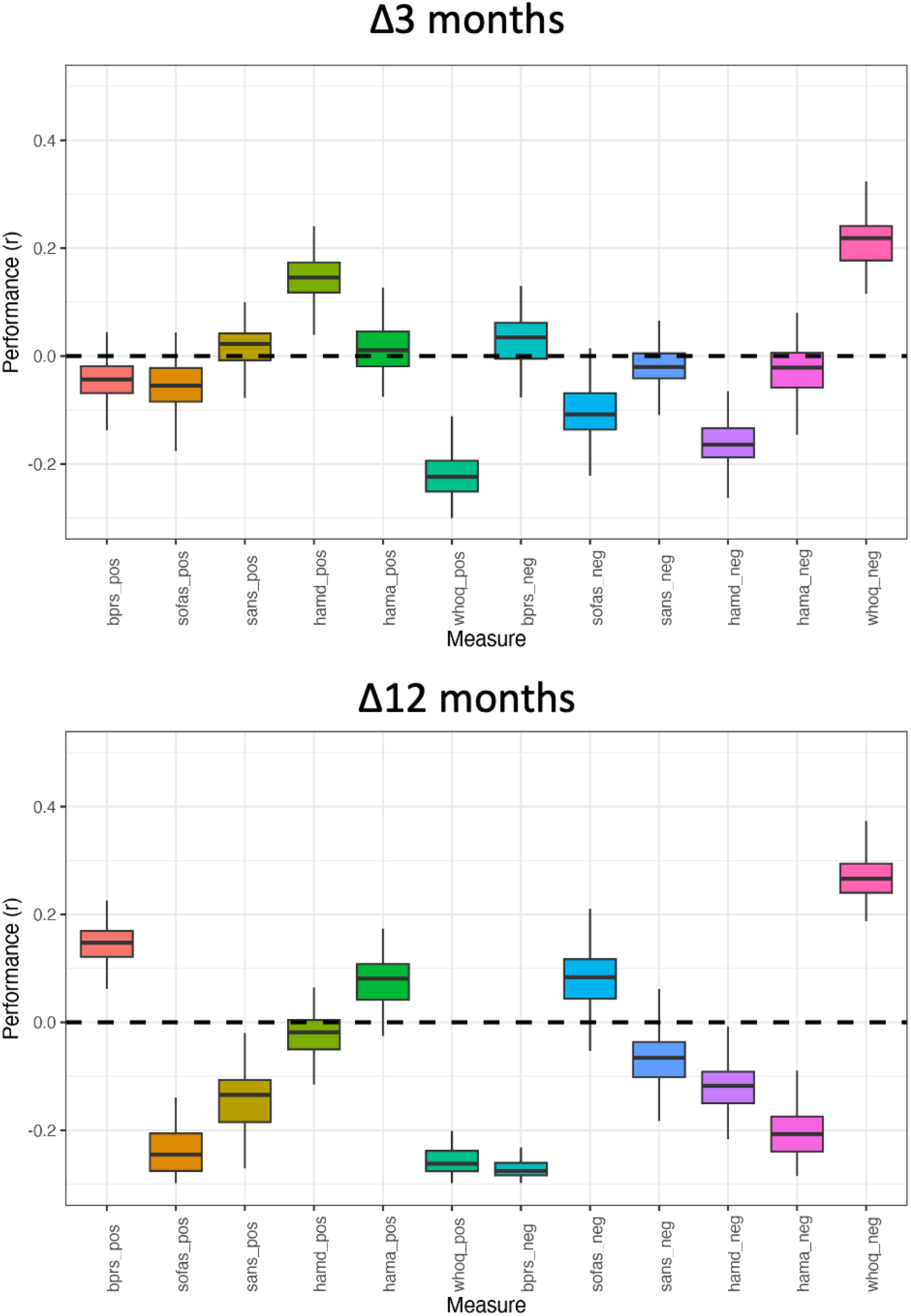
Product-moment correlation between observed and predicted behavioural change score (prediction performance) between baseline and 3-months (**top**) and baseline and 12-months (**bottom**) for the two primary outcome measures (Social and Occupational Functional Assessment Scale; SOFAS; Brief Psychiatric Rating Scale; BPRS), as well as the four secondary outcome measures (SANS; Scales for the Assessment of Negative Symptoms; HAMD; Hamilton Depression Rating Scale; HAMA; Hamilton Anxiety Rating Scale; WHO; World Health Organisation Quality of Life Scale) . For each of the scales, two models were computed, that selected connections that were either positively (pos) or negatively (neg) correlated with the outcome measure (see Methods). No p<.05 FDR significant associations were found for any scale. WHOQ at 12months was significant at p=.047, uncorrected.

**SFig6.**
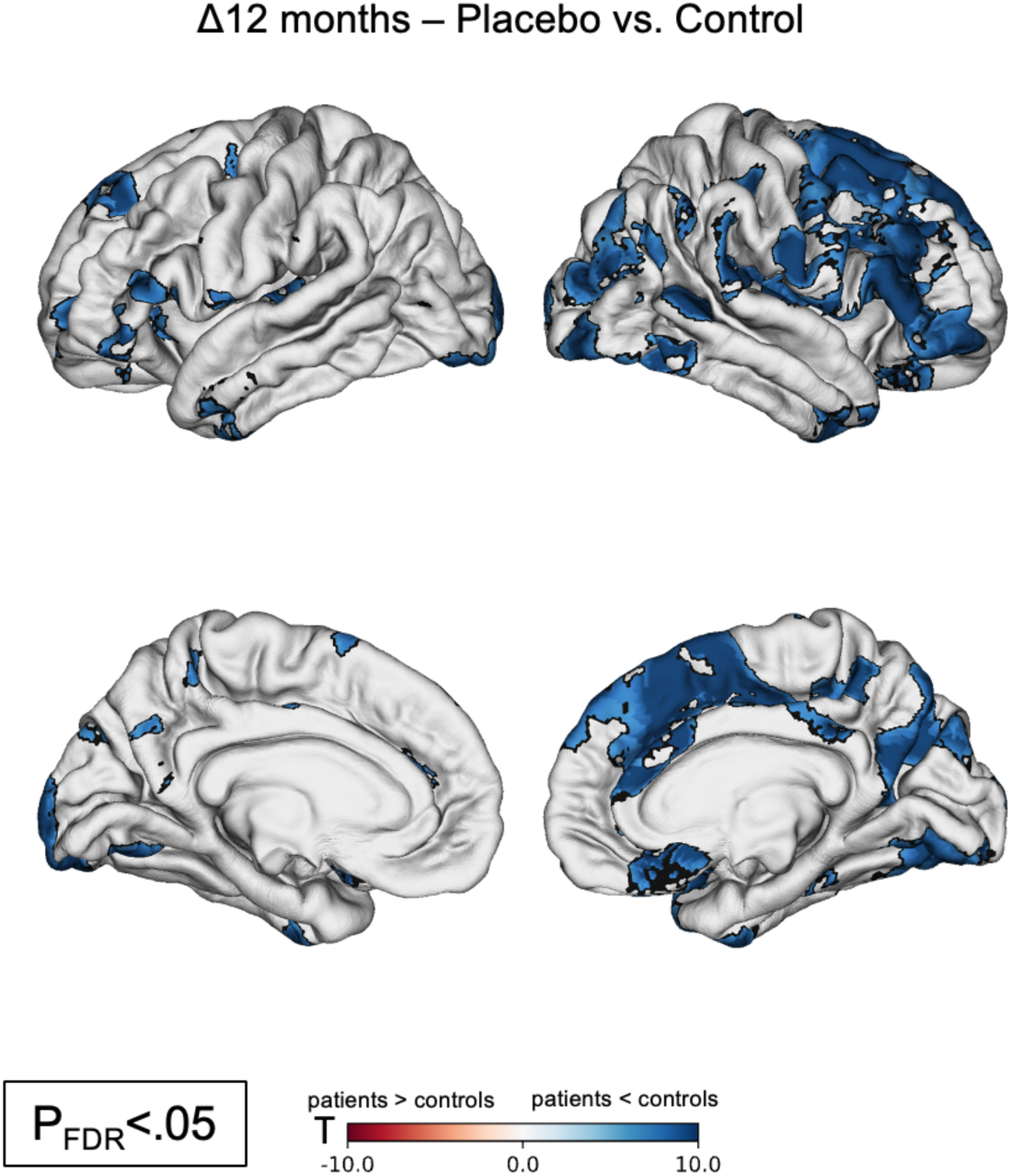
Illness-related cortical thinning from baseline to 12-months (p_FDR_<.05) remains largely unchanged when covarying for benzodiazepine exposure. All other contrasts remained non-significant.

**STable 1.**
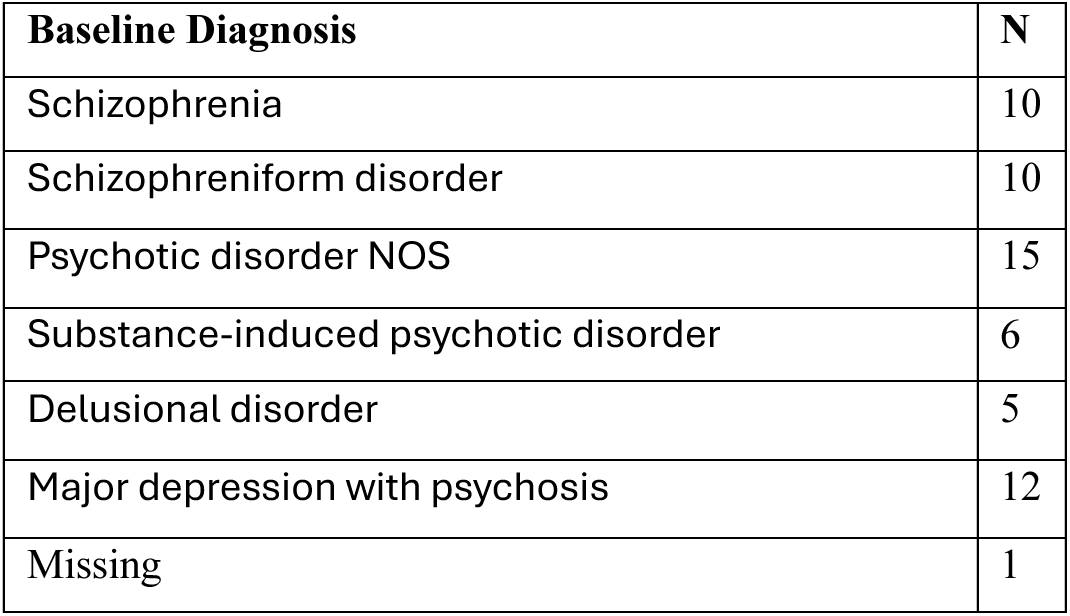

| **Baseline Diagnosis** | **N** |
| --- | --- |
| Schizophrenia | 10 |
| Schizophreniform disorder | 10 |
| Psychotic disorder NOS | 15 |
| Substance-induced psychotic disorder | 6 |
| Delusional disorder | 5 |
| Major depression with psychosis | 12 |
| Missing | 1 |

## References

1. Collaborators GMD. Global, regional, and national burden of 12 mental disorders in 204 countries and territories, 1990–2019: a systematic analysis for the Global Burden of Disease Study 2019. The Lancet Psychiatry 2022; 9(2): 137–50.

2. Ferrari AJ, Santomauro DF, Aali A, et al. Global incidence, prevalence, years lived with disability (YLDs), disability-adjusted life-years (DALYs), and healthy life expectancy (HALE) for 371 diseases and injuries in 204 countries and territories and 811 subnational locations, 1990–2021: a systematic analysis for the Global Burden of Disease Study 2021. The Lancet 2024; 403(10440): 2133–61.

3. Fusar-Poli P, Meyer-Lindenberg A. Forty years of structural imaging in psychosis: promises and truth. Acta Psychiatr Scand 2016; 134(3): 207–24.

4. van Haren NEM. Changes in Cortical Thickness During the Course of Illness in Schizophrenia. Archives of General Psychiatry 2011; 68(9): 871.

5. Andreasen NC, Nopoulos P, Magnotta V, Pierson R, Ziebell S, Ho B-C. Progressive Brain Change in Schizophrenia: A Prospective Longitudinal Study of First-Episode Schizophrenia. Biological psychiatry 2011; 70(7): 672–9.

6. Vita A, De Peri L, Deste G, Sacchetti E. Progressive loss of cortical gray matter in schizophrenia: a meta-analysis and meta-regression of longitudinal MRI studies. Translational Psychiatry 2012; 2(11): e190-e.

7. Konopaske GT, Dorph-Petersen K-A, Sweet RA, et al. Effect of chronic antipsychotic exposure on astrocyte and oligodendrocyte numbers in macaque monkeys. Biological Psychiatry 2008; 63(8): 759–65.

8. Konopaske GT, Dorph-Petersen K-A, Pierri JN, Wu Q, Sampson AR, Lewis DA. Effect of chronic exposure to antipsychotic medication on cell numbers in the parietal cortex of macaque monkeys. Neuropsychopharmacology: Official Publication of the American College of Neuropsychopharmacology 2007; 32(6): 1216–23.

9. Dorph-Petersen K-A, Pierri JN, Perel JM, Sun Z, Sampson AR, Lewis DA. The Influence of Chronic Exposure to Antipsychotic Medications on Brain Size before and after Tissue Fixation: A Comparison of Haloperidol and Olanzapine in Macaque Monkeys. Neuropsychopharmacology 2005; 30(9): 1649–61.

10. Vernon AC, Natesan S, Modo M, Kapur S. Effect of Chronic Antipsychotic Treatment on Brain Structure: A Serial Magnetic Resonance Imaging Study with Ex Vivo and Postmortem Confirmation. Biological Psychiatry 2011; 69(10): 936–44.

11. Veijola J, Guo JY, Moilanen JS, et al. Longitudinal Changes in Total Brain Volume in Schizophrenia: Relation to Symptom Severity, Cognition and Antipsychotic Medication. PLOS ONE 2014; 9(7): e101689.

12. Ho B-C, Andreasen NC, Ziebell S, Pierson R, Magnotta V. Long-term Antipsychotic Treatment and Brain Volumes. Archives of general psychiatry 2011; 68(2): 128–37.

13. Voineskos AN, Mulsant BH, Dickie EW, et al. Effects of Antipsychotic Medication on Brain Structure in Patients With Major Depressive Disorder and Psychotic Features: Neuroimaging Findings in the Context of a Randomized Placebo-Controlled Clinical Trial. JAMA Psychiatry 2020.

14. Fusar-Poli P, Smieskova R, Kempton MJ, Ho BC, Andreasen NC, Borgwardt S. Progressive brain changes in schizophrenia related to antipsychotic treatment? A meta-analysis of longitudinal MRI studies. Neuroscience and Biobehavioral Reviews 2013; 37(8): 1680–91.

15. Huhtaniska S, Jääskeläinen E, Hirvonen N, et al. Long-term antipsychotic use and brain changes in schizophrenia–a systematic review and meta-analysis. Human Psychopharmacology: Clinical and Experimental 2017; 32(2): e2574.

16. Haijma SV, Van Haren N, Cahn W, Koolschijn PCMP, Hulshoff Pol HE, Kahn RS. Brain Volumes in Schizophrenia: A Meta-Analysis in Over 18 000 Subjects. Schizophrenia Bulletin 2013; 39(5): 1129–38.

17. Chen AT, Nasrallah HA. Neuroprotective effects of the second generation antipsychotics. Schizophrenia Research 2019.

18. Lieberman JA, Tollefson GD, Charles C, et al. Antipsychotic Drug Effects on Brain Morphology in First-Episode Psychosis. Archives of General Psychiatry 2005; 62(4): 361–70.

19. Haddad PM, Correll CU. The acute efficacy of antipsychotics in schizophrenia: a review of recent meta-analyses. Therapeutic advances in psychopharmacology 2018; 8(11): 303–18.

20. Højlund M, Kemp AF, Haddad PM, Neill JC, Correll CU. Standard versus reduced dose of antipsychotics for relapse prevention in multi-episode schizophrenia: a systematic review and meta-analysis of randomised controlled trials. The Lancet Psychiatry 2021; 8(6): 471–86.

21. Correll CU, Solmi M, Croatto G, et al. Mortality in people with schizophrenia: a systematic review and meta−analysis of relative risk and aggravating or attenuating factors. World Psychiatry 2022; 21(2): 248–71.

22. Wunderink L, Nieboer RM, Wiersma D, Sytema S, Nienhuis FJ. Recovery in remitted first-episode psychosis at 7 years of follow-up of an early dose reduction/discontinuation or maintenance treatment strategy: long-term follow-up of a 2-year randomized clinical trial. JAMA psychiatry 2013; 70(9): 913–20.

23. Francey S, O’Donoghue B, Nelson B, et al. Psychosocial Intervention with or without Antipsychotic Medication for First Episode Psychosis: A Randomized Noninferiority Clinical Trial. Schizophrenia Bulletin Open 2020.

24. Chopra S, Fornito A, Francey SM, et al. Differentiating the effect of antipsychotic medication and illness on brain volume reductions in first-episode psychosis: A Longitudinal, Randomised, Triple-blind, Placebo-controlled MRI Study. Neuropsychopharmacology 2021; 46(8): 1494–501.

25. Fischl B, Dale AM. Measuring the thickness of the human cerebral cortex from magnetic resonance images. Proceedings of the National Academy of Sciences 2000; 97(20): 11050–5.

26. Clarkson MJ, Cardoso MJ, Ridgway GR, et al. A comparison of voxel and surface based cortical thickness estimation methods. Neuroimage 2011; 57(3): 856–65.

27. Panizzon MS, Fennema-Notestine C, Eyler LT, et al. Distinct genetic influences on cortical surface area and cortical thickness. Cerebral cortex 2009; 19(11): 2728–35.

28. Winkler AM, Kochunov P, Blangero J, et al. Cortical thickness or grey matter volume? The importance of selecting the phenotype for imaging genetics studies. Neuroimage 2010; 53(3): 1135–46.

29. Grasby KL, Jahanshad N, Painter JN, et al. The genetic architecture of the human cerebral cortex. Science 2020; 367(6484): eaay6690.

30. Vijayakumar N, Allen NB, Youssef G, et al. Brain development during adolescence: A mixed−longitudinal investigation of cortical thickness, surface area, and volume. Human brain mapping 2016; 37(6): 2027–38.

31. Bethlehem RAI, Seidlitz J, White SR, et al. Brain charts for the human lifespan. Nature 2022; 604(7906): 525–33.

32. Geschwind DH, Rakic P. Cortical evolution: judge the brain by its cover. Neuron 2013; 80(3): 633–47.

33. Francey SM, O’Donoghue B, Nelson B, et al. Psychosocial intervention with or without antipsychotic medication for first-episode psychosis: a randomized noninferiority clinical trial. Schizophrenia Bulletin Open 2020; 1(1): sgaa015.

34. O’Donoghue B, Francey SM, Nelson B, et al. Staged treatment and acceptability guidelines in early psychosis study (STAGES): A randomized placebo controlled trial of intensive psychosocial treatment plus or minus antipsychotic medication for first-episode psychosis with low-risk of self-harm or aggression. Study protocol and baseline characteristics of participants. Early Intervention in Psychiatry 2019; 0(0).

35. Fischl B. FreeSurfer. Neuroimage 2012; 62(2): 774–81.

36. Reuter M, Schmansky NJ, Rosas HD, Fischl B. Within-subject template estimation for unbiased longitudinal image analysis. Neuroimage 2012; 61(4): 1402–18.

37. Bernal-Rusiel JL, Reuter M, Greve DN, Fischl B, Sabuncu MR, Initiative AsDN. Spatiotemporal linear mixed effects modeling for the mass-univariate analysis of longitudinal neuroimage data. Neuroimage 2013; 81: 358–70.

38. Bernal-Rusiel JL, Greve DN, Reuter M, Fischl B, Sabuncu MR, Initiative AsDN. Statistical analysis of longitudinal neuroimage data with linear mixed effects models. Neuroimage 2013; 66: 249–60.

39. Benjamini Y, Krieger AM, Yekutieli D. Adaptive linear step-up procedures that control the false discovery rate. Biometrika 2006; 93(3): 491–507.

40. Chopra S, Francey SM, O’Donoghue B, et al. Functional connectivity in antipsychotic-treated and antipsychotic-naive patients with first-episode psychosis and low risk of self-harm or aggression: a secondary analysis of a randomized clinical trial. JAMA psychiatry 2021; 78(9): 994–1004.

41. Shen X, Finn ES, Scheinost D, et al. Using connectome-based predictive modeling to predict individual behavior from brain connectivity. nature protocols 2017; 12(3): 506–18.

42. Chopra S, Levi PT, Holmes A, et al. Brainwide Anatomical Connectivity and Prediction of Longitudinal Outcomes in Antipsychotic-Naïve First-Episode Psychosis. Biological Psychiatry 2025; 97(2): 157–66.

43. Francey S, O’Donoghue B, Nelson B, et al. Psychosocial Intervention With or Without Antipsychotic Medication for First-Episode Psychosis: A Randomized Noninferiority Clinical Trial. Schizophrenia Bulletin Open 2020; 1(sgaa015).

44. Yeo BT, Krienen FM, Sepulcre J, et al. The organization of the human cerebral cortex estimated by intrinsic functional connectivity. Journal of neurophysiology 2011.

45. Schaefer A, Kong R, Gordon EM, et al. Local-Global Parcellation of the Human Cerebral Cortex from Intrinsic Functional Connectivity MRI. Cerebral Cortex (New York, NY: 1991) 2018; 28(9): 3095–114.

46. von Economo CF, Koskinas GN. Die cytoarchitektonik der hirnrinde des erwachsenen menschen: J. Springer; 1925.

47. Scholtens LH, de Reus MA, de Lange SC, Schmidt R, van den Heuvel MP. An MRI Von Economo – Koskinas atlas. NeuroImage 2018; 170: 249–56.

48. Markello RD, Misic B. Comparing spatial null models for brain maps. NeuroImage 2021; 236: 118052.

49. Alexander-Bloch AF, Shou H, Liu S, et al. On testing for spatial correspondence between maps of human brain structure and function. Neuroimage 2018; 178: 540–51.

50. Markello RD, Hansen JY, Liu Z-Q, et al. Neuromaps: structural and functional interpretation of brain maps. Nature Methods 2022; 19(11): 1472–9.

51. Hawrylycz MJ, Lein ES, Guillozet-Bongaarts AL, et al. An anatomically comprehensive atlas of the adult human brain transcriptome. Nature 2012; 489(7416): 391–9.

52. Jorstad NL, Close J, Johansen N, et al. Transcriptomic cytoarchitecture reveals principles of human neocortex organization. Science 2023; 382(6667): eadf6812.

53. Zhang X-H, Anderson KM, Dong H-M, et al. The Cellular Underpinnings of the Human Cortical Connectome. bioRxiv 2023.

54. Jorstad NL, Song JH, Exposito-Alonso D, et al. Comparative transcriptomics reveals human-specific cortical features. Science 2023; 382(6667): eade9516.

55. Zhao Y, Zhang Q, Shah C, et al. Cortical thickness abnormalities at different stages of the illness course in schizophrenia: a systematic review and meta-analysis. JAMA psychiatry 2022; 79(6): 560–70.

56. Harrison PJ, Freemantle N, Geddes JR. Meta-analysis of brain weight in schizophrenia. Schizophrenia research 2003; 64(1): 25–34.

57. Akudjedu TN, Tronchin G, McInerney S, et al. Progression of neuroanatomical abnormalities after first-episode of psychosis: A 3-year longitudinal sMRI study. Journal of Psychiatric Research 2020; 130: 137–51.

58. Collins MA, Ji JL, Chung Y, et al. Accelerated cortical thinning precedes and predicts conversion to psychosis: The NAPLS3 longitudinal study of youth at clinical high-risk. Mol Psychiatry 2022.

59. Ziermans TB, Schothorst PF, Schnack HG, et al. Progressive Structural Brain Changes During Development of Psychosis. Schizophrenia Bulletin 2010; 38(3): 519–30.

60. Sun D, Stuart GW, Jenkinson M, et al. Brain surface contraction mapped in first-episode schizophrenia: a longitudinal magnetic resonance imaging study. Molecular Psychiatry 2009; 14(10): 976–86.

61. Cannon TD, Chung Y, He G, et al. Progressive reduction in cortical thickness as psychosis develops: a multisite longitudinal neuroimaging study of youth at elevated clinical risk. Biol Psychiatry 2015; 77(2): 147–57.

62. Sun D, Phillips L, Velakoulis D, et al. Progressive Brain Structural Changes Mapped as Psychosis Develops in ‘At Risk’ Individuals. Schizophrenia research 2009; 108(1-3): 85–92.

63. Guo S, Palaniyappan L, Liddle PF, Feng J. Dynamic cerebral reorganization in the pathophysiology of schizophrenia: a MRI-derived cortical thickness study. Psychological medicine 2016; 46(10): 2201–14.

64. Li M, Deng W, Li Y, et al. Ameliorative patterns of grey matter in patients with first-episode and treatment-naïve schizophrenia. Psychological Medicine 2023; 53(8): 3500–10.

65. Lewis DA, González-Burgos G. Neuroplasticity of neocortical circuits in schizophrenia. Neuropsychopharmacology 2008; 33(1): 141–65.

66. Lesh TA, Tanase C, Geib BR, et al. A Multimodal Analysis of Antipsychotic Effects on Brain Structure and Function in First-Episode Schizophrenia. JAMA Psychiatry 2015; 72(3): 226.

67. Gao X, Zhang W, Yao L, et al. Association between structural and functional brain alterations in drug-free patients with schizophrenia: a multimodal meta-analysis. Journal of Psychiatry and Neuroscience 2018; 43(2): 131–42.

68. Palaniyappan L. Clusters of psychosis: compensation as a contributor to the heterogeneity of schizophrenia. Journal of Psychiatry & Neuroscience: JPN 2023; 48(4): E325.

69. Ho B-C, Andreasen NC, Nopoulos P, Arndt S, Magnotta V, Flaum M. Progressive structural brain abnormalities and their relationship to clinical outcome: a longitudinal magnetic resonance imaging study early in schizophrenia. Archives of general Psychiatry 2003; 60(6): 585–94.

70. Berthet P, Haatveit BC, Kjelkenes R, et al. A 10-year longitudinal study of brain cortical thickness in people with first-episode psychosis using normative models. Schizophrenia Bulletin 2025; 51(1): 95–107.

71. James A, Javaloyes A, James S, Smith D. Evidence for non-progressive changes in adolescent-onset schizophrenia: follow-up magnetic resonance imaging study. The British Journal of Psychiatry 2002; 180(4): 339–44.

72. Palaniyappan L, Das TK, Winmill L, Hough M, James A, Palaniyappan L. Progressive post-onset reorganisation of MRI-derived cortical thickness in adolescents with schizophrenia. Schizophr Res 2019; 208: 477–8.

73. Haukvik UK, Hartberg CB, Nerland S, et al. No progressive brain changes during a 1-year follow-up of patients with first-episode psychosis. Psychological Medicine 2016; 46(3): 589–98.

74. Schaufelberger M, Lappin J, Duran F, et al. Lack of progression of brain abnormalities in first-episode psychosis: a longitudinal magnetic resonance imaging study. Psychological medicine 2011; 41(8): 1677–89.

75. Wen K, Zhao Y, Gong Q, et al. Cortical thickness abnormalities in patients with first episode psychosis: a meta-analysis of psychoradiologic studies and replication in an independent sample. Psychoradiology 2021; 1(4): 185–98.

76. Emsley R. Antipsychotics and structural brain changes: could treatment adherence explain the discrepant findings? Therapeutic Advances in Psychopharmacology 2023; 13: 20451253231195258.

77. Emsley R, du Plessis S, Phahladira L, et al. Antipsychotic treatment effects and structural MRI brain changes in schizophrenia. Psychological Medicine 2023; 53(5): 2050–9.

78. Bartzokis G, Lu PH, Amar CP, et al. Long acting injection versus oral risperidone in first-episode schizophrenia: differential impact on white matter myelination trajectory. Schizophrenia research 2011; 132(1): 35–41.

79. Tishler T, Ellingson B, Salvadore G, et al. Effect of treatment with paliperidone palmitate versus oral antipsychotics on frontal lobe intracortical myelin volume in participants with recent-onset schizophrenia: magnetic resonance imaging results from the DREaM study. Schizophrenia Research 2023; 255: 195–202.

80. Wang C, Tishler TA, Nuechterlein KH, Ellingson BM. Cortical thickness, gray-white matter contrast, and intracortical myelin in first-episode schizophrenia patients treated with long-acting paliperidone palmitate versus oral antipsychotics. Psychiatry Research 2023; 326: 115364.

81. Vita A, De Peri L, Deste G, Barlati S, Sacchetti E. The Effect of Antipsychotic Treatment on Cortical Gray Matter Changes in Schizophrenia: Does the Class Matter? A Meta-analysis and Meta-regression of Longitudinal Magnetic Resonance Imaging Studies. Biological Psychiatry 2015; 78(6): 403–12.

82. Ansell BRE, Dwyer DB, Wood SJ, et al. Divergent effects of first-generation and second-generation antipsychotics on cortical thickness in first-episode psychosis. Psychological Medicine 2015; 45(03): 515–27.

83. Shao Y, Peng H, Huang Q, Kong J, Xu H. Quetiapine mitigates the neuroinflammation and oligodendrocyte loss in the brain of C57BL/6 mouse following cuprizone exposure for one week. European Journal of Pharmacology 2015; 765: 249–57.

84. de Bartolomeis A, Barone A, Begni V, Riva MA. Present and future antipsychotic drugs: A systematic review of the putative mechanisms of action for efficacy and a critical appraisal under a translational perspective. Pharmacological research 2022; 176: 106078.

85. Ceraso A, Lin JJ, Schneider-Thoma J, et al. Maintenance treatment with antipsychotic drugs in schizophrenia: a Cochrane systematic review and meta-analysis. Schizophrenia Bulletin 2022; 48(4): 738–40.

86. Palomero-Gallagher N, Zilles K. Cortical layers: Cyto-, myelo-, receptor-and synaptic architecture in human cortical areas. Neuroimage 2019; 197: 716–41.

87. Baker JT, Holmes AJ, Masters GA, et al. Disruption of Cortical Association Networks in Schizophrenia and Psychotic Bipolar Disorder. JAMA psychiatry 2014; 71(2): 109–18.

88. Arumuham A, Nour MM, Veronese M, Onwordi EC, Rabiner EA, Howes OD. The histamine system and cognitive function: An in vivo H3 receptor PET imaging study in healthy volunteers and patients with schizophrenia. Journal of Psychopharmacology 2023; 37(10): 1011–22.

89. López-Figueroa AL, Norton CS, López-Figueroa MO, et al. Serotonin 5-HT1A, 5-HT1B, and 5-HT2A receptor mRNA expression in subjects with major depression, bipolar disorder, and schizophrenia. Biological psychiatry 2004; 55(3): 225–33.

90. Diez-Alarcia R, Muguruza C, Rivero G, et al. Opposite alterations of 5HT2A receptor brain density in subjects with schizophrenia: relevance of radiotracers pharmacological profile. Translational Psychiatry 2021; 11(1): 302.

91. Egerton A, Modinos G, Ferrera D, McGuire P. Neuroimaging studies of GABA in schizophrenia: a systematic review with meta-analysis. Translational psychiatry 2017; 7(6): e1147–e.

92. Duncan LE, Li T, Salem M, et al. Mapping the cellular etiology of schizophrenia and complex brain phenotypes. Nature Neuroscience 2025: 1–11.

93. Lewis DA, Curley AA, Glausier JR, Volk DW. Cortical parvalbumin interneurons and cognitive dysfunction in schizophrenia. Trends in neurosciences 2012; 35(1): 57–67.

94. Tayoshi SY, Nakataki M, Sumitani S, et al. GABA concentration in schizophrenia patients and the effects of antipsychotic medication: a proton magnetic resonance spectroscopy study. Schizophrenia research 2010; 117(1): 83–91.

95. Goto N, Yoshimura R, Kakeda S, et al. No alterations of brain GABA after 6 months of treatment with atypical antipsychotic drugs in early-stage first-episode schizophrenia. Progress in Neuro-Psychopharmacology and Biological Psychiatry 2010; 34(8): 1480–3.

96. Uliana DL, Lisboa JRF, Gomes FV, Grace AA. The excitatory-inhibitory balance as a target for the development of novel drugs to treat schizophrenia. Biochemical Pharmacology 2024: 116298.

97. Leucht S, Kane JM, Kissling W, Hamann J, Etschel E, Engel R. Clinical implications of Brief Psychiatric Rating Scale scores. British Journal of Psychiatry 2005; 187(4): 366–71.

